# Real-world Effectiveness of Sotrovimab for the Early Treatment of COVID-19 During SARS-CoV-2 Delta and Omicron Waves in the United States

**DOI:** 10.1101/2022.09.07.22279497

**Authors:** Mindy M. Cheng, Carolina Reyes, Sacha Satram, Helen Birch, Daniel C. Gibbons, Myriam Drysdale, Christopher F. Bell, Anvar Suyundikov, Xiao Ding, M. Cyrus Maher, Wendy Yeh, Amalio Telenti, Lawrence Corey

## Abstract

**Background:** Sotrovimab, a recombinant human monoclonal antibody (mAb) against SARS-CoV-2 had US FDA Emergency Use Authorization (EUA) for the treatment of high-risk outpatients with mild-to-moderate COVID-19 from May 26, 2021 to April 5, 2022. The study objective was to evaluate the real-world effectiveness of sotrovimab in reducing the risk of 30-day all-cause hospitalization and/or mortality during the time period when the prevalence of circulating SARS-CoV-2 variants was changing between Delta and Omicron sub-lineages in the US.

**Methods:** A retrospective analysis was conducted on de-identified claims data for 1,530,501 patients diagnosed with COVID-19 (ICD-10: U07.1) from September 1, 2021, to April 30, 2022, in the FAIR Health National Private Insurance Claims (FH NPIC^®^) database. Patients meeting EUA high-risk criteria were identified via pre-specified ICD-10-CM diagnoses in records ≤24 months prior to their first COVID-19 diagnosis and divided into two cohorts based on claimed procedural codes: treated with sotrovimab (“sotrovimab”) and not treated with a mAb (“no mAb”). All-cause hospitalizations and facility-reported all-cause mortality within 30 days of diagnosis (“30-day hospitalization or mortality”) were identified. Multivariable and propensity score-matched Poisson and logistic regressions were conducted to estimate the adjusted relative risk (RR) and odds of 30-day hospitalization or mortality among those treated with sotrovimab compared with those not treated with a mAb.

**Results:** Of the high-risk COVID-19 patients identified, 15,633 were treated with sotrovimab and 1,514,868 were not treated with a mAb. Compared with the no mAb cohort, the sotrovimab cohort was older and had a higher proportion of patients across the majority of high-risk conditions. In the no mAb cohort, 84,307 (5.57%) patients were hospitalized and 8,167 (0.54%) deaths were identified, while in the sotrovimab cohort, 418 (2.67%) patients were hospitalized and 13 (0.08%) deaths were identified. After adjusting for potential confounders, high-risk COVID-19 patients treated with sotrovimab had a 55% relative risk reduction of 30-day hospitalization or mortality (RR: 0.45, 95% CI: 0.41,0.49) and an 85% relative risk reduction of 30-day mortality (RR: 0.15, 95% CI: 0.08, 0.29) compared with high-risk patients not treated with a mAb. From September 2021 to April 2022, sotrovimab maintained clinical effectiveness with relative risk reductions of 30-day hospitalization or mortality ranging from 46% to 71%. Stratifying by high-risk condition, sotrovimab-treated patients exhibited statistically significant relative risk reductions of 30-day hospitalization or mortality compared with the no mAb cohort across all high-risk conditions (*P*<0.0001), ranging from 44% among pregnant women to 70% among patients 65 years and older.

**Conclusion:** In this large, US real-world, observational study of high-risk COVID-19 patients with reported diagnosis between September 2021 and April 2022 during the Delta and early Omicron variant waves, treatment with sotrovimab was associated with reduced risk of 30-day all-cause hospitalization and facility-reported mortality compared with no mAb treatment. Sotrovimab clinical effectiveness persisted throughout the months when Delta and early Omicron sub-lineages were the predominant circulating variants in the US, though there was an uncertain RR estimate in April 2022 with wide confidence intervals due to the small sample size. Sotrovimab clinical effectiveness also persisted among all high-risk subgroups assessed.

## INTRODUCTION

As of July 15, 2022, the severe acute respiratory syndrome coronavirus 2 (SARS-CoV-2) has caused over 557 million confirmed cases of the coronavirus disease (COVID-19) globally (WHO COVID-19 Dashboard). Although most cases of COVID-19 are mild to moderate, approximately 5-10% of cases are severe (Gavriatopoulou et al., 2021). It has been estimated that 18.2 million excess deaths worldwide were attributable to the COVID-19 pandemic over the course of 2020 and 2021 (COVID-19 Excess Mortality Collaborators).

Sotrovimab is a dual-action, recombinant human monoclonal antibody (mAb) that binds to a conserved epitope on the spike protein receptor binding domain of SARS-CoV-2 and has the ability to neutralize the virus and recruit the immune system to kill already infected cells *in vitro* and *in vivo* (Cathcart et al., 2021; Case et al., 2022). On May 26, 2021, the US Food and Drug Administration (FDA) issued an Emergency Use Authorization (EUA) for the use of sotrovimab (500 mg intravenous [IV]) for the treatment of mild-to-moderate COVID-19 in SARS-CoV-2 positive adults and pediatric patients (12 years of age or older weighing at least 40 kg) at high risk for severe COVID-19 (FDA News Release). The EUA for sotrovimab (500 mg IV) was issued based on interim safety and efficacy results from the phase 3, randomized, controlled clinical trial COMET-ICE (COVID-19 Monoclonal Antibody Efficacy Trial–Intent to Care Early), which showed that relative to placebo, sotrovimab was associated with an 85% (97.24% CI: 44-96%) relative risk reduction of all-cause hospitalization (lasting >24 hours) or death due to any cause within 29 days of treatment among high-risk outpatients (FDA News Release; Gupta et al., 2021). The final results of the COMET-ICE trial reported that 1% (6/528) of the sotrovimab-treated cohort had a 29-day hospitalization or mortality event compared with 6% (30/529) of the placebo cohort, resulting in a 79% (RR: 0.21, 95% CI: 0.09, 0.50) relative risk reduction in the same primary efficacy endpoint (Gupta et al., 2022).

The pre-clinical and clinical phases of sotrovimab (500 mg IV) development leading up to the EUA occurred prior to the SARS-CoV-2 Delta variant wave and when the population was mostly unvaccinated. Since the FDA EUA of sotrovimab, new SARS-CoV-2 variants of concern (VOC) have emerged, including Delta and Omicron sub-lineages. Although sotrovimab has retained activity against most known VOC *in vitro*, due to changes in *in vitro* potency relative to wild-type strain, the FDA deauthorized sotrovimab for the treatment of COVID-19 on a rolling basis across states with a >50% prevalence of Omicron BA.2; sotrovimab was deauthorized across the entire US on April 5, 2022 (FDA Fact Sheet).

Uncertainty remains regarding how *in vitro* antibody neutralization activity translates to clinical effectiveness, especially for dual-action antibodies like sotrovimab, which have potent effector function, including antibody-dependent cellular cytotoxicity (ADCC) and antibody-dependent cellular phagocytosis (ADCP) (Case et al., 2022). In order to assess real-world sotrovimab clinical effectiveness, we conducted a retrospective analysis of patients diagnosed with COVID-19 at high-risk of disease progression using a large, nationally representative insurance claims database. Specifically, the objectives of these analyses were to: 1) describe the demographics and clinical characteristics of patients treated with sotrovimab (500 mg IV) and not treated with a mAb; and 2) analyze the real-world clinical effectiveness of sotrovimab in reducing the risk of 30-day hospitalization and/or mortality among the overall treated cohort and among high-risk sub-groups from September 2021 to April 2022, during the SARS-CoV-2 Delta and early Omicron variant waves in the US.

## METHODS

A retrospective cohort analysis was conducted using the FAIR Health National Private Insurance Claims (FH NPIC^®^) database which is comprised of medical and dental claims submitted by over 70 private insurers across 50 US states, Puerto Rico, and the US Virgin Islands. The FH NPIC database undergoes rigorous internal validation processes and meets industry data security standards as demonstrated by its HITRUST, SOC 2, and CMS QE certifications. FAIR Health researchers were responsible for operationalizing all analyses upon study initiation in February 2022 pursuant to a data license and use agreement. Study sponsors did not have access to the FAIR Health database or to any patient-level data and were only provided with monthly reports (March through June 2022) comprised of de-identified, aggregated cohort level data in summary tables for patients with a claimed COVID-19 diagnosis date between September 1, 2021, and April 30, 2022.

The authors conducted a retrospective cohort study (secondary research) using de-identified and aggregated data licensed from a third party, FAIR Health, in compliance with 45 CFR 164.514(a)-(c) and the Health Insurance Portability and Accountability Act. Patient level identifiers were removed and were coded in such a way that the data could not be linked back to subjects from whom they were originally collected prior to the authors gaining access to it. This research, which used the de-identified licensed data described above, does not require IRB or ethics review, as analyses with these data do not meet the definition of ‘research involving human subjects’ as defined within 45 CFR 46.102(f) which stipulates human subjects as living individuals about whom an investigator obtains identifiable private information for research purposes.

### Study Population

The analysis included aggregated claims records for 1,530,501 de-identified patients in the FH NPIC database with a diagnosis of COVID-19 (ICD-10: U07.1) recorded between September 1, 2021, through April 30, 2022. The FH NPIC database is not linked to electronic health records; therefore, diagnoses, disease severity, and COVID-19 related hospitalizations or deaths could not be confirmed by laboratory test results or medical records.

Patients at high risk of disease progression were identified via pre-specified ICD-10-CM diagnosis codes aligned with sotrovimab’s EUA in the 24 months leading up to their first COVID-19 diagnosis date. Pre-specified high-risk conditions included: age (≥ 65 years), body mass index (BMI) ≥25 kg/m^2^, pregnancy, chronic kidney disease (CKD; any stage), type 1 or 2 diabetes, immunocompromising conditions (HIV, autoimmune disease, Hodgkin’s lymphoma, non-Hodgkin’s lymphoma, leukemia, solid cancer), immunosuppressive therapy (systemic corticosteroids and non-corticosteroid immunosuppressants billed using HCPCS [Healthcare Common Procedure Coding System] codes), chronic obstructive pulmonary disease (COPD), asthma, chronic lung disease, sickle cell disease, congenital heart disease, acquired heart disease, cardiovascular disease, hypertension, and neurodevelopmental disorders. Additionally, patients were considered high risk if pre-specified claims for a medical device (or components) outside of a non-acute care facility setting were identified, including non-invasive ventilation, oxygen therapy, renal replacement therapy, total parenteral nutrition, tracheostomy/other endotracheal airway, ventilation, ventricular assist, gastro- or jejunostomy, or Mitrofanoff catheter.

Individuals with missing data in claims records for age, gender, or geography were excluded from this analysis due to the inability to confirm unique identity and characterize the individual’s risk status.

### Study Variables

The diagnosis month refers to the month and year of the first COVID-19 diagnosis recorded in a patient’s claims record between September 1, 2021, and April 30, 2022. The diagnosis month category is a grouping of months that served as a proxy for the predominant circulating variant(s) or time period when a circulating variant became predominant in the US per the Centers for Disease Control and Prevention (CDC) COVID Data Tracker, Nowcast (CDC Nowcast). During the time period of September 1 through November 30, 2021, Delta was the predominant circulating variant in the US (>99% prevalence). During December 1, 2021, through February 28, 2022, Omicron BA.1 (and sub-lineages) became the predominant circulating variant, and during March 1 through April 30, 2022, Omicron BA.2 (and sub-lineages) became the predominant circulating variant.

Based on the state in which the COVID-19 diagnosis was recorded, patients were assigned to a US Department of Health and Human Services (HHS) designated region used by the CDC COVID Data Tracker, Nowcast, for variant monitoring and reporting: Region 1 (CT, ME, MA, NH, RI, VT), Region 2 (NJ, NY), Region 3 (DE, DC, MD, PA, VA, WV), Region 4 (AL, FL, GA, KY, MS, NC, SC, TN), Region 5 (IL, IN, MI, MN, OH, WI), Region 6 (AR, LA, NM, OK, TX), Region 7 (IA, KS, MO, NE), Region 8 (CO, MT, ND, SD, UT, WY), Region 9 (AZ, CA, HI, NV), and Region 10 (AK, ID, OR, WA).

The FH NPIC database included only COVID-19 vaccinations with claims submitted to a contributing insurer (“documented COVID-19 vaccine”). Those without a documented COVID-19 vaccine were of uncertain vaccination status as they could be unvaccinated or vaccinated without a submitted claim to a contributing insurer. The FH NPIC database captures limited data for oral drugs sold in the retail pharmacy setting or billed using National Drug Codes (NDC). Although antiviral therapy was present in the data, the variable was omitted from the analysis because of insufficient sample size for analysis (≤7 patients were identified in each cohort with a claim for an antiviral therapy after the COVID-19 diagnosis date).

### Exposure Variables

Claimed HCPCS codes for drug or infusion administration (in an outpatient setting) within 7 days of recorded diagnosis were used to divide high-risk patients into two cohorts: sotrovimab and not treated with a mAb authorized for early treatment of COVID-19 (“no mAb”). Supplementary Table 1 lists the HCPCS codes used to identify sotrovimab treatment, the mAbs screened to identify the no mAb cohort, and prophylaxis mAbs that were excluded from the analysis.

### Outcome Variables

Three outcomes of interest were assessed in this analysis: all-cause hospitalization within 30 days of claimed COVID-19 diagnosis (“hospitalization”), facility-reported all-cause mortality within 30 days of claimed diagnosis (“mortality”), and the composite outcome of all-cause hospitalization or facility-reported mortality within 30 days of claimed diagnosis date (“30-day hospitalization or mortality”). All-cause hospitalizations were identified in claims by Bill Types with beginning digits 11*, 12*, and 18*, corresponding to hospital facility inpatient care. Intensive care unit (ICU)-related hospitalizations were identified by Revenue Codes: 0200, 0202, 0203, 0206, 0208, 0209. Deaths were identified from claims records based on the discharge status reported by healthcare facilities for which a billable medical service was provided. No information regarding cause of death was available in the data.

### Statistical Analyses

All statistical analyses were performed using SAS software, version 9.4 (SAS Institute Inc., Cary, North Carolina). Continuous variables were summarized by mean, standard deviation (SD) and median, interquartile range (IQR). The chi-square test for categorical variables and the t-test for continuous variables were performed to compare statistical differences by treatment cohort. *P*-values were not adjusted for multiplicity. All statistical comparisons were two-sided, with significance assumed at α ≤ 0.05.

Multivariable Poisson and logistic regression analyses were conducted to assess the impact of sotrovimab (versus no mAb) on 30-day hospitalization and/or mortality, adjusting for potential confounders. Confounders were selected from demographic and clinical characteristics, based on *a priori* evidence that these factors are associated with receipt of treatment or risk of hospitalization and/or death (CDC Assessing Risk). Covariates in the Poisson and logistic regression models were the same and included diagnosis month category, region, gender, age, rurality (urban/rural based on geozip where COVID-19 diagnosis was recorded), obesity (BMI ≥ 30 kg/m^2^), pregnancy, CKD, diabetes, immunocompromising conditions (includes immunosuppressive therapy), lung disease (includes COPD, asthma, chronic lung disease), cardiovascular disease (includes acquired heart disease, congenital heart disease, cardiovascular disease, hypertension), medical device, and documented COVID-19 vaccine. Adjusted relative risk (RR) and 95% confidence intervals (CI) were derived from the Poisson regression model with robust error variances and presented as primary results in this manuscript. Adjusted odds ratios (OR) and 95% CIs were estimated from the logistic regression model and are presented in the Supplementary Tables. For analyses stratified by diagnosis month, high-risk conditions, and documented COVID-19 vaccine, the multivariable Poisson and logistic regression models were run separately for each sub-group.

As a sensitivity analysis for the comparison between the sotrovimab and no mAb cohorts, propensity score (PS)-matched Poisson and logistic regression analyses were also conducted to estimate the RR (95% CI) and OR (95% CI), respectively, of 30-day hospitalization or mortality among PS-matched cohorts. The effect of the propensity score matching was to balance the treatment groups to reduce potential bias associated with treatment selection. Multinomial logistic regression was used to calculate the propensity score once across the entire cohort—the conditional probability that each patient would be assigned to a specific treatment group given that patient’s pretreatment variables. The treatment cohorts were matched on diagnosis month, region, gender, age, rurality, obesity, pregnancy, CKD, diabetes, immunocompromising conditions (including immunosuppressive therapy), lung disease (including asthma, COPD, chronic lung disease), cardiovascular disease (including heart disease and hypertension) and medical device. Greedy nearest neighbor matching with a caliper of 0.2 and matching ratio of 1:4 (1 sotrovimab treated patient was matched to 4 no mAb patients) without replacement was performed. The PS match was successful as all the covariates included in the propensity score model had a standardized mean difference (SMD) of ≤ 0.10 between the sotrovimab and no mAb cohorts.

## RESULTS

### Cohort Characteristics

As shown in Table 1, among 1,530,501 high-risk patients diagnosed with COVID-19 between September 1, 2021, through April 30, 2022, 15,633 were treated with sotrovimab and 1,514,868 were not treated with a mAb. Compared with the no mAb cohort, the sotrovimab cohort was older (median age [years]: 55 versus 48; *P<*0.0001) and had more elderly patients (20% vs 13% ≥ 65 years; *P* <0.0001). Patients in both cohorts were more likely to have been diagnosed with COVID-19 between December 1, 2021, and February 28, 2022, the time period when Omicron BA.1 became the predominant circulating variant (CDC Nowcast). Approximately 20% of the sotrovimab cohort had at least one claim for a COVID-19 vaccine compared with approximately 15% of the no mAb cohort. Hypertension was the most common high-risk condition among both cohorts. Compared with the no mAb cohort, the sotrovimab cohort had a higher proportion of patients across each of the high-risk conditions. Notably, approximately 42% of the sotrovimab cohort had a diagnosis of an immunocompromising condition and/or immunosuppressive therapy compared with approximately 25% of the no mAb cohort.

**Table 1:**
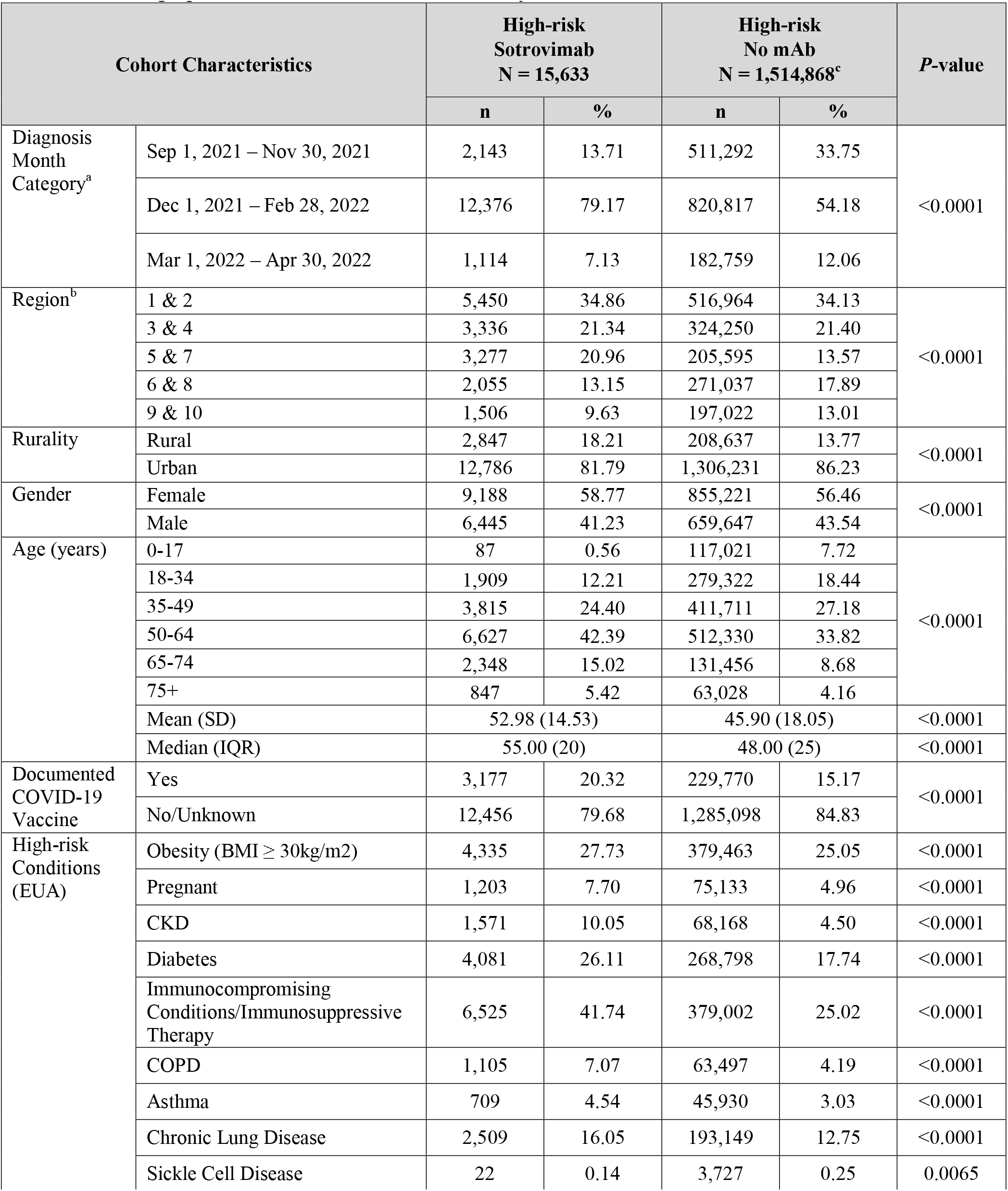
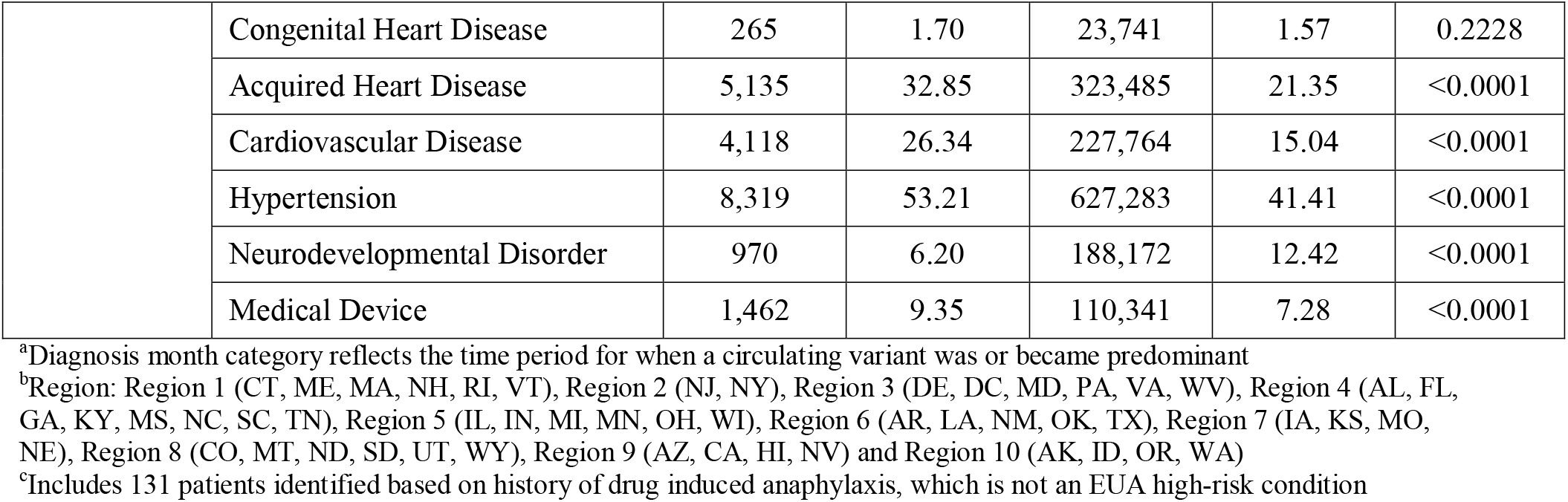
Demographic and Clinical Characteristics by Treatment Cohort

| Cohort Characteristics |  | High-risk<br>Sotrovimab<br>N = 15,633 |  | High-risk<br>No mAb<br>N = 1,514,868<sup>c</sup> |  | P-value |
| --- | --- | --- | --- | --- | --- | --- |
|  |  | n | % | n | % |  |
| Diagnosis Month Category<sup>a</sup> | Sep 1, 2021 – Nov 30, 2021 | 2,143 | 13.71 | 511,292 | 33.75 | <0.0001 |
|  | Dec 1, 2021 – Feb 28, 2022 | 12,376 | 79.17 | 820,817 | 54.18 |  |
|  | Mar 1, 2022 – Apr 30, 2022 | 1,114 | 7.13 | 182,759 | 12.06 |  |
| Region<sup>b</sup> | 1 & 2 | 5,450 | 34.86 | 516,964 | 34.13 | <0.0001 |
|  | 3 & 4 | 3,336 | 21.34 | 324,250 | 21.40 |  |
|  | 5 & 7 | 3,277 | 20.96 | 205,595 | 13.57 |  |
|  | 6 & 8 | 2,055 | 13.15 | 271,037 | 17.89 |  |
|  | 9 & 10 | 1,506 | 9.63 | 197,022 | 13.01 |  |
| Rurality | Rural | 2,847 | 18.21 | 208,637 | 13.77 | <0.0001 |
|  | Urban | 12,786 | 81.79 | 1,306,231 | 86.23 |  |
| Gender | Female | 9,188 | 58.77 | 855,221 | 56.46 | <0.0001 |
|  | Male | 6,445 | 41.23 | 659,647 | 43.54 |  |
| Age (years) | 0-17 | 87 | 0.56 | 117,021 | 7.72 | <0.0001 |
|  | 18-34 | 1,909 | 12.21 | 279,322 | 18.44 |  |
|  | 35-49 | 3,815 | 24.40 | 411,711 | 27.18 |  |
|  | 50-64 | 6,627 | 42.39 | 512,330 | 33.82 |  |
|  | 65-74 | 2,348 | 15.02 | 131,456 | 8.68 |  |
|  | 75+ | 847 | 5.42 | 63,028 | 4.16 |  |
|  | Mean (SD) | 52.98 (14.53) |  | 45.90 (18.05) |  | <0.0001 |
|  | Median (IQR) | 55.00 (20) |  | 48.00 (25) |  | <0.0001 |
| Documented COVID-19 Vaccine | Yes | 3,177 | 20.32 | 229,770 | 15.17 | <0.0001 |
|  | No/Unknown | 12,456 | 79.68 | 1,285,098 | 84.83 |  |
| High-risk Conditions (EUA) | Obesity (BMI ≥ 30kg/m<sup>2</sup>) | 4,335 | 27.73 | 379,463 | 25.05 | <0.0001 |
|  | Pregnant | 1,203 | 7.70 | 75,133 | 4.96 | <0.0001 |
|  | CKD | 1,571 | 10.05 | 68,168 | 4.50 | <0.0001 |
|  | Diabetes | 4,081 | 26.11 | 268,798 | 17.74 | <0.0001 |
|  | Immunocompromising Conditions/Immunosuppressive Therapy | 6,525 | 41.74 | 379,002 | 25.02 | <0.0001 |
|  | COPD | 1,105 | 7.07 | 63,497 | 4.19 | <0.0001 |
|  | Asthma | 709 | 4.54 | 45,930 | 3.03 | <0.0001 |
|  | Chronic Lung Disease | 2,509 | 16.05 | 193,149 | 12.75 | <0.0001 |
|  | Sickle Cell Disease | 22 | 0.14 | 3,727 | 0.25 | 0.0065 |
|  | Congenital Heart Disease | 265 | 1.70 | 23,741 | 1.57 | 0.2228 |
|  | Acquired Heart Disease | 5,135 | 32.85 | 323,485 | 21.35 | <0.0001 |
|  | Cardiovascular Disease | 4,118 | 26.34 | 227,764 | 15.04 | <0.0001 |
|  | Hypertension | 8,319 | 53.21 | 627,283 | 41.41 | <0.0001 |
|  | Neurodevelopmental Disorder | 970 | 6.20 | 188,172 | 12.42 | <0.0001 |
|  | Medical Device | 1,462 | 9.35 | 110,341 | 7.28 | <0.0001 |

<sup>a</sup>Diagnosis month category reflects the time period for when a circulating variant was or became predominant
<sup>b</sup>Region: Region 1 (CT, ME, MA, NH, RI, VT), Region 2 (NJ, NY), Region 3 (DE, DC, MD, PA, VA, WV), Region 4 (AL, FL, GA, KY, MS, NC, SC, TN), Region 5 (IL, IN, MI, MN, OH, WI), Region 6 (AR, LA, NM, OK, TX), Region 7 (IA, KS, MO, NE), Region 8 (CO, MT, ND, SD, UT, WY), Region 9 (AZ, CA, HI, NV) and Region 10 (AK, ID, OR, WA)
<sup>c</sup>Includes 131 patients identified based on history of drug induced anaphylaxis, which is not an EUA high-risk condition

### 30-day All-Cause Hospitalization and Facility-Reported Mortality

Of the 15,633 high-risk COVID-19 patients treated with sotrovimab, 418 (2.67%) patients were hospitalized within 30 days of diagnosis (Table 2), of whom 65 (15.55%) were admitted to the ICU. Of the 1,514,868 high-risk patients in the no mAb cohort, 84,307 (5.57%) were hospitalized within 30 days of diagnosis, of whom 24,489 (29.05%) had an ICU admission. A total of 13 (0.08%) facility-reported all-cause deaths were identified within 30 days of diagnosis in the sotrovimab cohort, with a mean time between reported diagnosis and death of 24 days. Within the no mAb cohort, 8,167 (0.54%) deaths were identified within 30 days of diagnosis, with a mean time between reported diagnosis and death of 21.9 days.

**Table 2:**
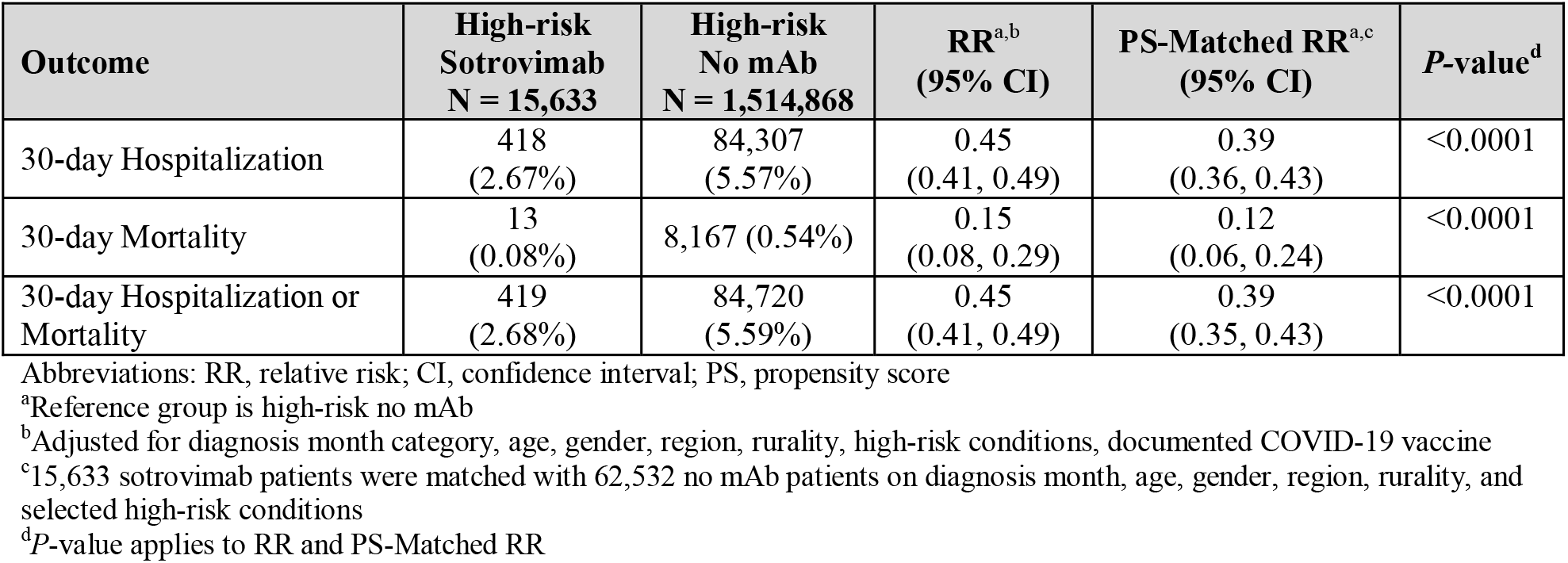
Adjusted and Propensity Score-Matched Risk of 30-day All-Cause Hospitalization or Facility-Reported Mortality

| Outcome | High-risk<br>Sotrovimab<br>N = 15,633 | High-risk<br>No mAb<br>N = 1,514,868 | RR<sup>a,b</sup><br>(95% CI) | PS-Matched RR<sup>a,c</sup><br>(95% CI) | P-value<sup>d</sup> |
| --- | --- | --- | --- | --- | --- |
| 30-day Hospitalization | 418<br>(2.67%) | 84,307<br>(5.57%) | 0.45<br>(0.41, 0.49) | 0.39<br>(0.36, 0.43) | <0.0001 |
| 30-day Mortality | 13<br>(0.08%) | 8,167 (0.54%) | 0.15<br>(0.08, 0.29) | 0.12<br>(0.06, 0.24) | <0.0001 |
| 30-day Hospitalization or<br>Mortality | 419<br>(2.68%) | 84,720<br>(5.59%) | 0.45<br>(0.41, 0.49) | 0.39<br>(0.35, 0.43) | <0.0001 |

Abbreviations: RR, relative risk; CI, confidence interval; PS, propensity score
<sup>a</sup>Reference group is high-risk no mAb
<sup>b</sup>Adjusted for diagnosis month category, age, gender, region, rurality, high-risk conditions, documented COVID-19 vaccine
<sup>c</sup>15,633 sotrovimab patients were matched with 62,532 no mAb patients on diagnosis month, age, gender, region, rurality, and selected high-risk conditions
<sup>d</sup>P-value applies to RR and PS-Matched RR

Among the 418 hospitalizations in the sotrovimab cohort, 61 (14.6%) cases had a reported primary cause of admission related to childbirth (labor and delivery), compared with 4,172 (4.9%) cases of the 84,307 hospitalizations identified in the no mAb cohort. Birth outcomes were not available in the data.

### Relative Risk of 30-day All-Cause Hospitalization or Facility-Reported Mortality

After adjusting for all other covariates in the Poisson regression model (Table 2), treatment with sotrovimab was associated with a 55% relative risk reduction of 30-day hospitalization (RR: 0.45, 95% CI: 0.41, 0.49), an 85% relative risk reduction of 30-day mortality (RR: 0.15, 95% CI: 0.08, 0.29), and a 55% relative risk reduction of 30-day hospitalization or mortality (RR: 0.45, 95% CI: 0.41, 0.49). Supplementary Table 2 presents ORs and 95% CIs of the full multivariable logistic regression model of factors predicting the odds of 30-day hospitalization or mortality. Increasing age, male gender, or having any of the high-risk conditions increased the odds of 30-day hospitalization or mortality.

A total of 15,633 high-risk patients treated with sotrovimab were PS-matched with 62,532 high-risk patients not treated with a mAb. PS matching successfully eliminated statistically significant differences in matched characteristics between treatment cohorts (Supplementary Table 3). Sotrovimab effectiveness was higher in the PS-matched analysis with a 61% relative risk reduction (RR: 0.39, 95% CI: 0.35, 0.43) of 30-day hospitalization or mortality and an 88% relative risk reduction (RR: 0.12, 95% CI: 0.06, 0.24) in mortality among sotrovimab treated patients versus patients not treated with a mAb. Odds ratios and 95% CIs for 30-day hospitalization or mortality between treatment cohorts are shown in Supplementary Table 4.

**Table 3:** Adjusted and Propensity Score-Matched Relative Risk of 30-Day All-Cause Hospitalization or Facility-Reported Mortality by Diagnosis Month

| Diagnosis Month | High-risk Sotrovimab |  | High-risk No mAb |  | RR<sup>b,c</sup><br>(95% CI) | PS-Matched RR<sup>b,d</sup><br>(95% CI) | P-value |
| --- | --- | --- | --- | --- | --- | --- | --- |
|  | n | %<sup>a</sup> | n | %<sup>a</sup> |  |  |  |
| September 2021 | 460 | 4.78% | 307,096 | 9.53% | 0.48<br>(0.32, 0.72) | 0.39<br>(0.26, 0.60) | 0.0002 |
| October 2021 | 451 | 2.66% | 84,367 | 8.62% | 0.29<br>(0.17, 0.51) | 0.27<br>(0.15, 0.47) | <0.0001 |
| November 2021 | 1,232 | 2.76% | 119,829 | 7.81% | 0.33<br>(0.23, 0.45) | 0.32<br>(0.22, 0.45) | <0.0001 |
| December 2021 | 5,188 | 3.14% | 455,706 | 3.90% | 0.49<br>(0.43, 0.57) | 0.45<br>(0.39, 0.53) | <0.0001 |
| January 2022 | 4,859 | 1.85% | 261,765 | 3.37% | 0.36<br>(0.30, 0.44) | 0.36<br>(0.29, 0.44) | <0.0001 |
| February 2022 | 2,329 | 3.26% | 103,346 | 6.90% | 0.43<br>(0.34, 0.54) | 0.40<br>(0.32, 0.50) | <0.0001 |
| March 2022 | 1,046 | 2.01% | 65,521 | 4.37% | 0.41<br>(0.27, 0.62) | 0.36<br>(0.23, 0.56) | <0.0001 |
| April 2022 | 68 | 1.47% | 117,238 | 1.90% | 0.54<br>(0.08, 3.54) | 0.32<br>(0.04, 2.38) | 0.5185 |

Abbreviations: RR, relative risk; CI, confidence interval; PS, propensity score
<sup>a</sup>The reported % is the number hospitalized or died in diagnosis month in treatment cohort / number in diagnosis month in treatment cohort
<sup>b</sup>Reference group is high-risk no mAb
<sup>c</sup>Adjusted for diagnosis month category, age, gender, region, rurality, high-risk conditions, and documented COVID-19 vaccine
<sup>d</sup>15,633 sotrovimab patients were matched with 62,532 no mAb patients on diagnosis month, age, gender, region, rurality, and selected high-risk conditions
<sup>e</sup>P-value applies to RR and PS-Matched RR

### Relative Risk of 30-day All-Cause Hospitalization or Facility-Reported Mortality by Diagnosis Month

As shown in Table 3, in the no mAb cohort, rates of 30-day hospitalization or mortality were lower in December 2021 through April 2022, when Omicron sub-lineages became the predominant circulating variants, compared with September through November 2021 when Delta was the predominant variant (Figure 1). From September 2021 to March 2022, treatment with sotrovimab was associated with statistically significant relative risk reductions of 30-day hospitalization or mortality ranging from 51% to 71% (PS-matched relative risk reduction from 55% to 73%). In April 2022, treatment with sotrovimab was associated with a non-significant 46% relative risk reduction (68% PS-matched relative risk reduction) of 30-day hospitalization or mortality. Figure 1 graphically represents the PS-matched relative risk of 30-day hospitalization or mortality for the sotrovimab cohort versus the no mAb cohort over time. In the upper panel of the figure, the monthly average US prevalence of the circulating variants is shown over the study time-period. The estimated RR for sotrovimab was found to be similar across three pandemic waves. Sotrovimab effectiveness was maintained as the average US prevalence of BA.2 and its sub-lineages increased to approximately 50% in March 2022. In April of 2022, the relative prevalence of BA.2 and its sub-lineages increased to 94% (CDC Nowcast). During the same time period, sotrovimab use had been largely discontinued, resulting in an uncertain RR estimate in April with wide confidence intervals most likely due to the small sample size. Odds ratios and 95% CIs for 30-day hospitalization or mortality between treatment cohorts by month of diagnosis are shown in Supplementary Table 5.

**Figure 1:**
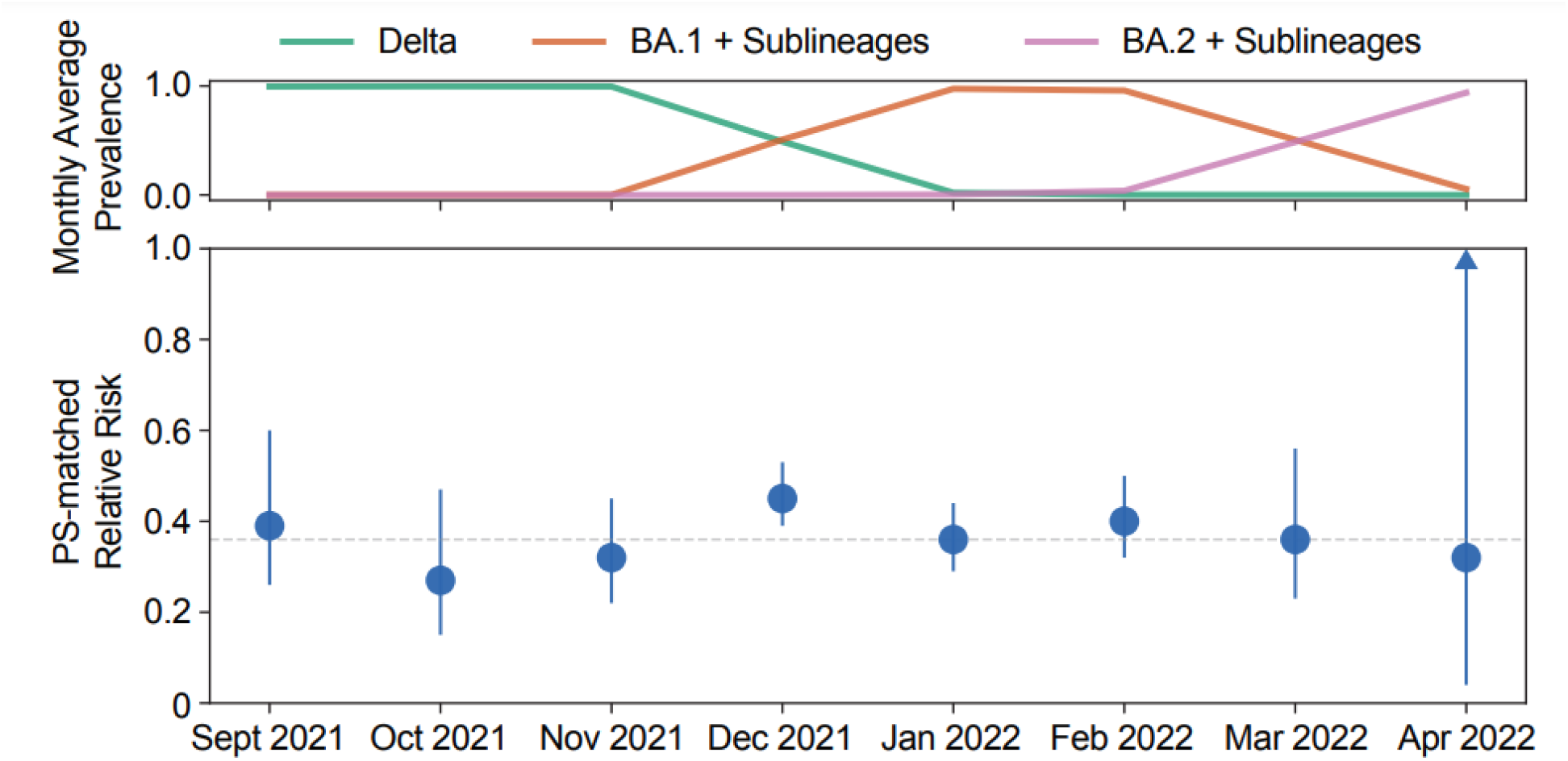
Propensity Score-Matched Relative Risk of 30-day All-Cause Hospitalization or Facility-Reported Mortality and COVID-19 Variant Prevalence Over Time. Monthly US average prevalences of Delta (green), BA.1 + sub-lineages (orange) and BA.2 + sub-lineages (pink) are depicted in the upper panel based on data from the Global Initiative on Sharing All Influenza Data (GISAID) (Elbe and Buckland-Merrett, 2017) and not necessarily representative of the study population. The lower panel shows the relative risk by month for the high-risk sotrovimab cohort relative to the high-risk no mAb cohort. Patients were propensity score matched on diagnosis month, age, gender, region, rurality, and selected high-risk conditions. Note: The 95% CI for the PS-matched RR for April 2022 is (0.04, 2.38)

### Relative Risk of 30-Day All-Cause Hospitalization or Facility-Reported Mortality by High-Risk Condition

Stratifying the multivariate regression model by high-risk condition (Figure 2), sotrovimab treated patients showed statistically significant relative risk reductions of 30-day hospitalization or mortality compared with patients in the no mAb cohort, ranging from 44% among pregnant women to 70% among patients 65 years and older. The subgroup of patients with an immunocompromising condition and/or immunosuppressive therapy had a 53% relative risk reduction (RR: 0.47, 95% CI: 0.41, 0.54) of 30-day hospitalization or mortality.

**Figure 2:**
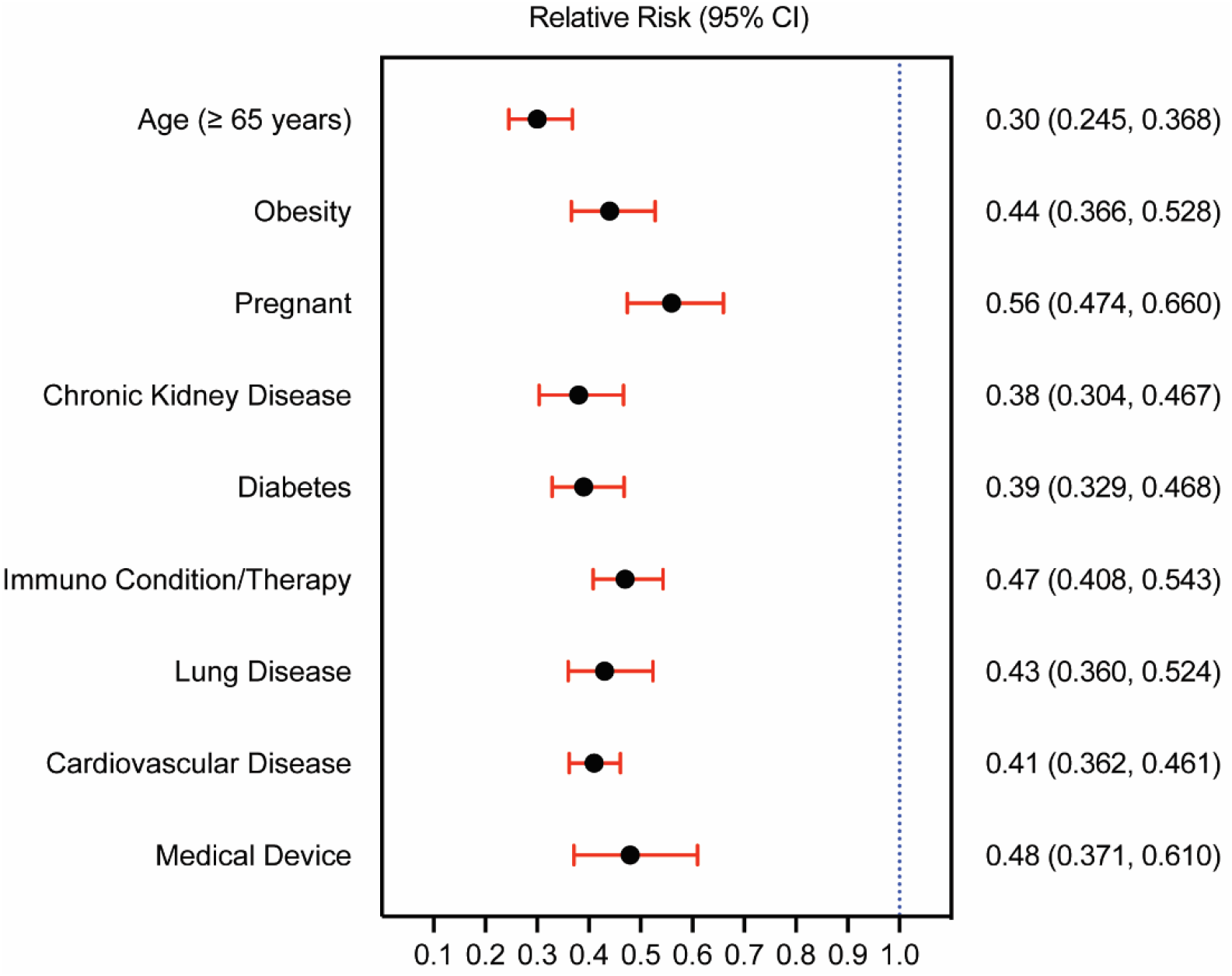
Relative Risk of 30-day All-Cause Hospitalization or Facility-Reported Mortality by High-Risk Condition. Adjusted relative risk for the sotrovimab cohort versus the no mAb cohort Note: *P*-value < 0.0001 for all high-risk conditions

### Impact of Documented COVID-19 Vaccine

Of the 3,177 patients with at least one documented COVID-19 vaccine in the sotrovimab cohort, 59 (1.86%) were hospitalized within 30 days of diagnosis. In the no mAb cohort, 229,770 patients had at least one documented COVID-19 vaccine, of whom 7,030 (3.06%) were hospitalized within 30 days of diagnosis. Among those with at least one documented COVID-19 vaccine, treatment with sotrovimab was associated with a 57% relative risk reduction (RR: 0.43, 95% CI: 0.34, 0.56) of 30-day hospitalization or mortality.

## DISCUSSION

In this real-world, retrospective claims database analysis of high-risk COVID-19 patients with reported diagnosis between September 1, 2021 through April 30, 2022, during the Delta and early Omicron waves, treatment with sotrovimab was associated with a 55% relative risk reduction (59% lower odds) of 30-day hospitalization or mortality and an 85% relative risk reduction and lower odds of facility-reported mortality compared with not being treated with a mAb. In the PS-matched analysis, the estimate of sotrovimab effectiveness was higher, with a 61% relative risk reduction (63% lower odds) of 30-day all-cause hospitalization or mortality and an 88% relative risk reduction and lower odds of mortality among sotrovimab treated patients versus patients not treated with a mAb, comparable to the COMET-ICE trial results (RR: 0.21, 95% CI: 0.09, 0.50). Across the months of the study from September 2021 to March 2022, sotrovimab maintained clinical effectiveness with relative risk reductions of 30-day hospitalization or mortality ranging from 51% to 71% (55% to 74% lower odds). In April 2022, treatment with sotrovimab was associated with a non-significant 46% relative risk reduction (48% lower odds) of 30-day hospitalization or mortality. Sotrovimab had been deauthorized across the entire US on April 5, 2022, leading to a small sample size (n=68) in the sotrovimab treated group. This small sample size, together with mixed variant prevalence and lack of specific sequencing data, limits our ability to draw conclusions about sotrovimab effectiveness in April 2022.

Approximately 42% of the sotrovimab cohort versus 25% of the no mAb cohort had a diagnosis of an immunocompromising condition and/or immunosuppressive therapy recorded within 24 months leading up to their COVID-19 diagnosis. It is well-documented that the immunocompromised/immunosuppressed population is at a higher risk of severe disease due to challenges in mounting robust antibody responses to the COVID-19 vaccine (Lee, et al., 2022; Parker et al., 2022). Sotrovimab was shown to be protective, with a statistically significant 53% relative risk reduction (57% lower odds) of 30-day hospitalization or mortality in this subgroup of immunocompromised/immunosuppressed patients. Furthermore, sotrovimab treatment was associated with relative risk reductions of 30-day hospitalization or mortality across all high-risk conditions examined, including a 70% lower risk among patients 65 years and older when compared with the no mAb cohort. Although the sotrovimab cohort had a higher rate (15%) of 30-day hospitalizations related to childbirth (labor and delivery) compared with the no mAb cohort (5%), after adjustment for confounders, sotrovimab conferred a statistically significant 44% relative risk reduction of 30-day hospitalization or mortality among pregnant women. However, maternal COVID-19 disease severity, the timing of sotrovimab treatment relative to labor and delivery, and birth outcomes could not be assessed in the database.

Approximately 20% of the sotrovimab cohort had at least one claim for a COVID-19 vaccine compared with approximately 15% of the no mAb cohort. However, it cannot be determined whether these patients were partially or fully vaccinated. The vaccination status of those without a documented COVID-19 vaccine in claims records is uncertain and is more likely reflective of limited billing for COVID-19 vaccinations by vaccine providers given US Government procurement and distribution, rather than the actual vaccination rates within the study population. Despite this uncertainty in vaccination status, sotrovimab treatment was associated with a 57% relative risk reduction of 30-day hospitalization or mortality among those with at least one documented COVID-19 vaccine compared with no mAb treatment. This estimate is consistent with the overall treated population estimate and may suggest that sotrovimab provides treatment benefits in the presence of COVID-19 vaccine antibodies. A large proportion of the study cohort was immunocompromised, and it is not clear to what extent this relationship with vaccine status was impacted by the large proportion of immunocompromised patients in the current study. To further elucidate this relationship, additional research is required among vaccinated patients within high-risk subgroups.

Aggarwal and colleagues used a similar approach to assess sotrovimab effectiveness among high-risk outpatients diagnosed with COVID-19 in Colorado during the period of October 1, 2021, through December 11, 2021, when Delta was the predominant circulating variant. Among 522 sotrovimab-treated patients PS-matched with 1,563 patients not treated with a mAb, sotrovimab treatment was associated with a 63% (OR: 0.37; 95% CI: 0.19, 0.66) lower odds of 28-day all-cause hospitalization and an 89% (OR: 0.11; 95% CI: 0.0, 0.79) lower odds of 28-day all-cause mortality, similar to our findings during the time period when Delta was the predominant variant (Aggarwal et al., 2022a). A subsequent study using similar methods assessed sotrovimab effectiveness among 5,205 PS-matched high-risk patients between December 26, 2021, and March 10, 2022, and found non-significant lower odds of 28-day all-cause hospitalization (OR: 0.82, 95% CI: 0.55, 1.19) and 28-day all-cause mortality (OR: 0.62, 95% CI: 0.07, 2.78) during the Omicron BA.1 wave (Aggarwal, et al., 2022b). In contrast, our findings showed that sotrovimab maintained statistically significant lower odds of 30-day all-cause hospitalization or mortality between 55%-67% during the December 2021 through March 2022 time period when Omicron BA.1 became the predominant circulating variant. Among high-risk subgroups, Aggarwal, et al. observed trending towards a lower odds of 28-day hospitalization among patients age ≥65 years (OR: 0.52, 95% CI: 0.30, 0.92), immunocompromised patients (OR: 0.63, 95% CI: 0.38, 1.04), and those with 2 or more comorbid conditions (OR: 0.65, 95% CI: 0.42, 1.01) who were treated with sotrovimab compared with those not treated with a mAb. The non-significant findings from Aggarwal, et al. may be due to the smaller sample size and lower power to detect differences between treatment groups (Aggarwal, et al., 2022b).

Finally, another small study in Qatar conducted during the reported Omicron BA.2 wave found non-significant higher odds (OR: 2.67, 95% CI: 0.60, 11.91) of progression to severe, critical, or fatal COVID-19 in sotrovimab treated patients compared with those untreated. In a subset of patients at higher risk of severe forms of COVID-19, the study additionally found non-significant higher odds (OR: 1.33, 95% CI: 0.44, 4.05) of progression to severe, critical, or fatal COVID-19 in sotrovimab treated patients compared with those untreated (Zaquot et al., 2022). However, there are some important differences in the study methodology that preclude an appropriate comparison to the current study findings, most notably, patients were excluded from the control group if they showed signs or symptoms of severe COVID-19 within 7 days of diagnosis. By doing so, this likely biased the sotrovimab treated group to be sicker and at a higher risk of disease progression compared with the control group, which may explain the observed results.

Over the course of 2021 and the first half of 2022, the COVID-19 pandemic has proceeded with SARS-CoV-2 Delta and Omicron (BA.1, BA.2, BA.3, BA.4, BA.5) VOC, with no clear end in sight (Telenti et al., 2022). The rapid rate of SARS-CoV-2 mutations makes it challenging to assess the effectiveness of interventions against emerging VOCs, especially when confounded by population immunity from vaccination or natural infection which may lower the mAb titers needed for neutralization in humans. Recent evidence suggests that *in vitro* neutralization activity may only be a partial determinant of sotrovimab efficacy, and that Fc-mediated effector functions such as ADCC and ADCP may contribute additional antiviral effects against SARS-CoV-2 Omicron variants based on recent *in vivo* studies in mice (Case et al., 2022). Effector function may explain the notable lack of correlation between *in vitro* neutralization and *in vivo* activity in animal studies and is a possible explanation for the preserved clinical effectiveness observed over time, even as BA.2 became more prevalent (Case et al., 2022). As such, real-world evidence obtained from usual clinical practice has become increasingly important to assess treatment effectiveness outside of *in vitro* and *in vivo* studies or tightly controlled clinical trials among a highly selected group of patients.

### Strengths Ȧ Limitations

The FH NPIC claims database is a useful resource to examine therapeutic strategies in actual clinical practice settings. To our knowledge, with over 1.5 million patients, our study is one of the largest to assess sotrovimab effectiveness among high-risk COVID-19 patients with geographic representation across the US. However, the primary purpose of insurance claims data is for billing and reimbursement as opposed to research. Therefore, the use of claims data limits the ability to verify the diagnostic conditions reported, disease severity, or treatment pathways. The data are also limited in capturing vaccinations (or unbilled services) and oral therapies dispensed through retail pharmacy or billed using NDC (eg, oral non-corticosteroids). Further, the FH NPIC database is comprised of healthcare claims only from private insurers in the US and does not include populations covered by public programs (eg, Medicare, Medicaid) or individuals without health insurance. However, given that the population identified in this study was patients at high-risk of severe disease who are more likely to utilize healthcare services, we expect the findings to be generalizable to other high-risk subpopulations. Facility-reported mortality likely underestimated the true number of deaths, as deaths were only identifiable from the discharge status reported by a healthcare facility for which a billable medical service was provided.

Finally, we cannot exclude the possibility of residual confounding due to the absence of race/ethnicity and variant sequencing data in the dataset. We used the month of COVID-19 diagnosis as a proxy for the likelihood of a given case of COVID-19 being attributable to the Delta, Omicron BA.1 or BA.2 variants. Circulating variants may influence clinicians’ decisions to treat and/or the specific regimen to administer, and this proxy variable may not adequately assess variant exposure for all patients in our study. We also adopted several sensitivity approaches to account for the effect of changing COVID-19 variants on treatment effectiveness, including propensity score matching as well as stratified Poisson and logistic regression analyses. Regardless of how we approached the analyses, receipt of sotrovimab among high-risk patients showed consistent protective benefits of preventing 30-day hospitalization or mortality. Further research is warranted utilizing sequencing data to confirm our study findings.

Patients in the no mAb cohort appeared to be eligible for sotrovimab per EUA criteria but were untreated for unknown reasons. If we applied the adjusted RR reductions in hospitalizations and mortality with receipt of sotrovimab to the no mAb cohort, an estimated 46,000 (43,000 – 50,000) hospitalizations and 7,000 (5,800 -7,500) deaths may have been avoided throughout the time period in this study. Reducing the disparity of non-clinical factors in the receipt of effective COVID-19 therapies will help improve outcomes among high-risk patients.

## CONCLUSIONS

In this large, US real-world, observational study of high-risk COVID-19 patients during the Delta and early Omicron waves, treatment with sotrovimab was associated with reduced risk of 30-day all-cause hospitalization or facility-reported mortality compared with no mAb treatment. Sotrovimab clinical effectiveness persisted throughout the months when Delta and early Omicron sub-lineages were the predominant circulating strains in the US, though there was an uncertain RR estimate in April 2022 with a wide confidence interval due to the small sample size. Sotrovimab clinical effectiveness also persisted among all high-risk subgroups assessed. It is imperative that interventions with demonstrated real-world clinical effectiveness be accessible to patients at high-risk of severe COVID-19 disease.

## Data Availability

N/A

## Funding

This study was funded by Vir Biotechnology and GSK.

## Acknowledgments

We thank Ali Russo, Alexander Mizenko, and Randi Scott from FAIR Health for conducting the data extraction, data analyses and statistical analyses for this study. Research for this study was based upon the data compiled and maintained by FAIR Health, Inc. The authors are solely responsible for the research and conclusions reflected in this article. FAIR Health is not responsible for any of the conclusions or opinions expressed herein.

We thank Wenjie Wang, Hong Wang, Rajendra Joshi, and Yimeng Lu for assistance with the visual presentation of the data. We thank Tamara Palagashvili and Don Hoang for publication support.

Editorial assistance was provided by Lumanity Scientific Inc. and was funded by Vir Biotechnology.

## Notes

### Competing Interest Statement

Mindy M. Cheng, Carolina Reyes, Sacha Satram, Anvar Suyundikov, Xiao Ding, M. Cyrus Maher, Wendy Yeh, and Amalio Telenti are employees of and shareholders of Vir Biotechnology. Helen Birch, Daniel C. Gibbons, Myriam Drysdale, and Christopher F. Bell are employees of and shareholders of GSK. Lawrence Corey has no conflicts of interest to disclose.

### Funding Statement

Study is sponsored by Vir Biotechnology in collaboration with GSK

