## Supplemental Data for "Real-world Effectiveness of Sotrovimab for the Early Treatment of COVID-19 During SARS-CoV-2 Delta and Omicron Waves in the United States"

**SUPPLEMENTARY TABLES**

Supplementary Table 1: HCPCS codes used to identify treatment with mAbs for early treatment and pre-exposure prophylaxis

| **Procedure Code** | **Description** |
| --- | --- |
| **Sotrovimab** | |
| M0247 | Intravenous infusion, sotrovimab, includes infusion and post administration monitoring |
| M0248 | Intravenous infusion, sotrovimab, includes infusion and post administration monitoring (home administration) |
| Q0247 | Injection, sotrovimab, 500 mg |
| **mAbs authorized for early treatment of COVID-19** | |
| M0240 | Intravenous infusion or subcutaneous injection, casirivimab and imdevimab |
| M0241 | Intravenous infusion or subcutaneous injection, casirivimab and imdevimab |
| M0243 | Intravenous infusion or subcutaneous injection, casirivimab and imdevimab |
| M0244 | Intravenous infusion or subcutaneous injection, casirivimab and imdevimab |
| M0245 | Intravenous infusion, bamlanivimab and etesevimab, includes infusion and post admin |
| M0246 | Intravenous infusion, bamlanivimab and etesevimab, includes infusion and post admin |
| M0239 | Intravenous infusion, bamlanivimab |
| M0222 | Intravenous injection, bebtelovimab, includes injection and post administration monitoring |
| M0223 | Intravenous injection, bebtelovimab, includes injection and post administration monitoring in the home or residence |
| Q0240 | Injection, casirivimab and imdevimab, 600 mg |
| Q0243 | Injection, casirivimab and imdevimab, 2400 mg |
| Q0244 | Injection, casirivimab and imdevimab, 1200 mg |
| Q0245 | Injection, bamlanivimab and etesevimab, 2100 mg |
| Q0239 | Injection, bamlanivimab 700 mg |
| Q0222 | Injection, bebtelovimab, 175 mg |
| **mAbs for pre-exposure prophylaxis** | |
| M0220 | Tixagevimab and cilgavimab injection |
| M0221 | Tixagevimab and cilgavimab injection, home administration |
| Q0220 | Tixagevimab and cilgavimab injection |
| Q0221 | Tixagevimab and cilgavimab, 600 mg |

Supplementary Table 2: Multivariable Logistic Regression of Factors Predicting the Risk of 30-day Hospitalization and Mortality

| **Covariates** | **OR** | **95% CI** |
| --- | --- | --- |
| Treatment | 0.41 | 0.37, 0.45 |
| No mAb (ref) |  |  |
| Sotrovimab | 0.41 | 0.37, 0.45 |
| Age (years) |  |  |
| 18-34 (ref) |  |  |
| 0-17 | 0.52 | 0.50, 0.55 |
| 35-49 | 1.23 | 1.20, 1.26 |
| 50-64 | 1.94 | 1.89, 2.00 |
| 65-74 | 2.91 | 2.82, 3.00 |
| 75+ | 4.92 | 4.75, 5.10 |
| Gender |  |  |
| Female (ref) |  |  |
| Male | 1.37 | 1.35, 1.40 |
| Diagnosis Date Category^a^ |  |  |
| Sep 1 – Nov 30, 2021(ref) |  |  |
| Dec 1, 2021 - Feb 28, 2022 | 0.45 | 0.41, 0.45 |
| Mar 1 – Apr 30, 2022 | 0.30 | 0.29, 0.31 |
| Region^b^ |  |  |
| 1 & 2 (ref) |  |  |
| 3 & 4 | 1.32 | 1.29, 1.34 |
| 5 & 7 | 1.52 | 1.48, 1.55 |
| 6 & 8 | 1.35 | 1.33, 1.38 |
| 9 & 10 | 1.30 | 1.27, 1.33 |
| Rurality |  |  |
| Urban (ref) |  |  |
| Rural | 0.97 | 0.96, 0.99 |
| No Documented COVID-19 Vaccination | 1.81 | 1.76, 1.85 |
| High Risk Conditions |  |  |
| Pregnant | 12.56 | 12.22, 12.91 |
| Obesity (BMI ≥ 30 kg/m^2^) | 1.15 | 1.13, 1.17 |
| Diabetes (Type 1 or 2) | 1.44 | 1.41, 1.46 |
| Cardiovascular/ Heart Disease/Hypertension | 1.39 | 1.37, 1.42 |
| Immunocompromising Conditions/Immunosuppressive Therapy | 1.16 | 1.14, 1.18 |
| CKD | 1.93 | 1.88, 1.98 |
| Lung Disease | 1.16 | 1.14, 1.19 |
| Medical Device | 1.23 | 1.20, 1.25 |

Abbreviations: OR, odds ratio; CI, confidence interval; BMI, body mass index

^a^Diagnosis Month Category reflects the time period in which a circulating variant became predominant; Sep-Nov 2021 (Delta), Dec 2021-Feb 2022 (BA.1), Mar-Apr 2022 (BA.2)

^b^Region: Region 1 (CT, ME, MA, NH, RI, VT), Region 2 (NJ, NY), Region 3 (DE, DC, MD, PA, VA, WV), Region 4 (AL, FL, GA, KY, MS, NC, SC, TN), Region 5 (IL, IN, MI, MN, OH, WI), Region 6 (AR, LA, NM, OK, TX), Region 7 (IA, KS, MO, NE), Region 8 (CO, MT, ND, SD, UT, WY), Region 9 (AZ, CA, HI, NV) and Region 10 (AK, ID, OR, WA)

Supplementary Table 3: Demographic and Clinical Characteristics by Treatment Cohort after Propensity Score Matching

| **Cohort Characteristics** | | **High-risk**  **Sotrovimab**  **N = 15,633** | | **High-risk**  **No mAb**  **N = 62,532** | | ***P-*value** |
| --- | --- | --- | --- | --- | --- | --- |
|  |  | **n** | **%** | **n** | **%** |  |
| Diagnosis Month Category | September 2021 | 460 | 2.94 | 1,897 | 3.03 | 0.8777 |
|  | October 2021 | 451 | 2.88 | 1,854 | 2.96 |  |
|  | November 2021 | 1,232 | 7.88 | 5,010 | 8.01 |  |
|  | December 2021 | 5,188 | 33.19 | 20,643 | 33.01 |  |
|  | January 2022 | 4,859 | 31.08 | 19,192 | 30.69 |  |
|  | February 2022 | 2,329 | 14.90 | 9,548 | 15.27 |  |
|  | March 2022 | 1,046 | 6.69 | 4,108 | 6.57 |  |
|  | April 2022 | 68 | 0.43 | 280 | 0.45 |  |
| Region | 1 & 2 | 5,450 | 34.86 | 21,881 | 34.99 | 0.9411 |
|  | 3 & 4 | 3,336 | 21.34 | 13.479 | 21.56 |  |
|  | 5 & 7 | 3,277 | 20.96 | 13,002 | 20.79 |  |
|  | 6 & 8 | 2,055 | 13.15 | 8,114 | 12.98 |  |
|  | 9 & 10 | 1,515 | 9.69 | 6,056 | 9.68 |  |
| Rurality | Rural | 2,847 | 18.21 | 11,173 | 17.87 | 0.3163 |
|  | Urban | 12,786 | 81.79 | 51,359 | 82.13 |  |
| Gender | Female | 9,188 | 58.77 | 36,708 | 58.70 | 0.8773 |
|  | Male | 6,445 | 41.23 | 25,824 | 41.30 |  |
| Age (years) | 0-17 | 87 | 0.56 | 293 | 0.47 | 0.5091 |
|  | 18-34 | 1,909 | 12.21 | 7,400 | 11.83 |  |
|  | 35-49 | 3,815 | 24.40 | 15,190 | 24.29 |  |
|  | 50-64 | 6,627 | 42.39 | 26,728 | 42.74 |  |
|  | 65-74 | 2,348 | 15.02 | 9,467 | 15.14 |  |
|  | 75+ | 847 | 5.42 | 3,454 | 5.52 |  |
| High-risk Conditions (EUA) | Obesity (BMI ≥ 30 kg/m^2^) | 4,335 | 27.73 | 17,411 | 27.84 | 0.7800 |
|  | Pregnant | 1,202 | 7.69 | 4,857 | 7.77 | 0.7508 |
|  | CKD | 1,571 | 10.05 | 5,881 | 9.40 | 0.0148 |
|  | Diabetes | 4,081 | 26.11 | 16,350 | 26.15 | 0.9190 |
|  | ImmunocompromisingConditions / Immunosuppressive Therapy | 6,525 | 41.74 | 26,087 | 41.72 | 0.9638 |
|  | Lung Disease | 3,546 | 22.68 | 14,017 | 22.42 | 0.4730 |
|  | Cardiovascular / Heart Disease / Hypertension | 9,856 | 63.05 | 39,559 | 63.26 | 0.6166 |
|  | Medical Device | 1,450 | 9.28 | 5,496 | 8.79 | 0.0573 |

Supplementary Table 4: Adjusted and Propensity Score Matched Logistic Regression of 30-day All-Cause Hospitalization or Facility-Reported Mortality

| **Outcome** | **High-risk**  **Sotrovimab**  **N=15,633** | **High-risk**  **No mAb**  **N=1,514,868** | **OR** ^a,b^  **(95% CI)** | **PS-Matched OR** ^a,c^  **(95% CI)** |
| --- | --- | --- | --- | --- |
| 30-day Hospitalization | 418  (2.67%) | 84,307 (5.57%) | 0.41  (0.37, 0.46) | 0.37 (0.34, 0.41) |
| 30-day Mortality | 13  (0.08%) | 8,167 (0.54%) | 0.15  (0.08, 0.29) | 0.12 (0.06, 0.23) |
| 30-day Hospitalization or Mortality | 419  (2.68%) | 84,720  (5.59%) | 0.41  (0.37, 0.45) | 0.37  (0.34, 0.41) |

Abbreviations: OR, odds ratio; CI, confidence interval; PS, propensity score

^a^Reference group is high-risk no mAb

^b^Adjusted for diagnosis month category, age, gender, region, rurality, high-risk conditions, documented COVID-19 vaccine

^c^15,633 sotrovimab patients were matched with 62,532 no mAb patients on diagnosis month, age, gender, region, rurality, and selected high-risk conditions

Supplementary Table 5: Adjusted and Propensity Score-Matched Logistic Regression of 30-Day All-Cause Hospitalization or Facility-Reported Mortality by Diagnosis Month

| **Diagnosis Month^a^** | **High-risk Sotrovimab** | | **High-risk No mAb** | | **OR**^b,c^  **(95% CI)** | **PS-Matched OR**^b,d^  **(95% CI)** |
| --- | --- | --- | --- | --- | --- | --- |
|  | n | % **^a^** | n | % **^a^** |  |  |
| September 2021 | 460 | 4.78% | 307,096 | 9.53% | 0.44  (0.28, 0.68) | 0.36  (0.23, 0.57) |
| October 2021 | 451 | 2.66% | 84,367 | 8.62% | 0.26  (0.15, 0.47) | 0.25  (0.14, 0.45) |
| November 2021 | 1,232 | 2.76% | 119,829 | 7.81% | 0.29  (0.21, 0.41) | 0.30  (0.21, 0.42) |
| December 2021 | 5,188 | 3.14% | 455,706 | 3.90% | 0.45  (0.39, 0.53) | 0.43  (0.37, 0.51) |
| January 2022 | 4,859 | 1.85% | 261,765 | 3.37% | 0.33  (0.27, 0.41) | 0.35  (0.28, 0.43) |
| February 2022 | 2,329 | 3.26% | 103,346 | 6.90% | 0.39 (0.31, 0.50) | 0.38  (0.30, 0.48) |
| March 2022 | 1,046 | 2.01% | 65,521 | 4.37% | 0.38  (0.24, 0.58) | 0.34  (0.22, 0.54) |
| April 2022 | 68 | 1.47% | 117,238 | 1.90% | 0.52 (0.07, 3.85) | 0.31  (0.04, 2.39) |

Abbreviations: OR, odds ratio; CI, confidence interval; PS, propensity score

^a^The reported % is the number nospitalized or died in diagnosis month in treatment cohort / number in diagnosis month in treatment cohort

^b^Reference group is high-risk no mAb

^c^Adjusted for diagnosis month category, age, gender, region, rurality, high-risk conditions, and documented COVID-19 vaccine

^d^15,633 sotrovimab patients were matched with 62,532 no mAb patients on diagnosis month, age, gender, region, rurality, and selected high-risk conditions
